# Transmission dynamics of Norovirus GII and Enterovirus in Switzerland during the COVID-19 pandemic (2021-2022) as evidenced in wastewater

**DOI:** 10.1101/2025.04.22.25326216

**Authors:** Jana S. Huisman, Shotaro Torii, Htet Kyi Wynn, Charles Gan, Irene K. Voellmy, Michael Huber, Timothy R. Julian, Tamar Kohn

**Affiliations:** Physics of Living Systems, Massachusetts Institute of Technology, Cambridge, MA, USA; Department of Urban Engineering, School of Engineering, University of Tokyo, Tokyo, Japan; Laboratory of Environmental Virology, School of Architecture, Civil and Environmental Engineering (ENAC), Ecole Polytechnique Fédérale de Lausanne (EPFL), Lausanne, Switzerland; Eawag, Swiss Federal Institute of Aquatic Science and Technology, Duebendorf, Switzerland; Institute of Medical Virology, University of Zurich, Zurich, Switzerland; Swiss Tropical and Public Health Institute, Allschwil, Switzerland; University of Basel, Basel, Switzerland

**Keywords:** effective reproductive number, wastewater based epidemiology, pathogen surveillance, norovirus, enterovirus

## Abstract

Noroviruses and enteroviruses are major causes of endemic gastrointestinal disease associated with substantial disease burden. However, viral gastroenteritis is often diagnosed based on symptoms, with etiology infrequently tested or reported, so little information exists on community-level transmission dynamics. In this study, we demonstrate that norovirus (NoV) genogroup II and enterovirus (EV) viral loads in wastewater reveal transmission dynamics of these viruses. We report NoV and EV concentrations in wastewater from 363 samples between December 5 2020 and October 10 2022 (sampled every second day). Virus concentrations in wastewater were low during 2021, and increased in 2022. Wastewater recapitulated periods of increased clinical cases, and also identified silent waves of transmission. We used the measured wastewater loads to estimate the effective reproductive number (Re). The Re for both NoV and EV peaked between 1.1-1.2. However, the usual seasonality of NoV transmission was upended by non-pharmaceutical interventions implemented to mitigate the COVID-19 pandemic, leading to correlated transmission dynamics of NoV GII and EV during 2021-2022. This highlights the use of wastewater to understand transmission dynamics of endemic enteric viruses and estimate relevant epidemiological parameters, including Re.

## Introduction

Human norovirus (NoV) and enterovirus (EV) are two major genera of enteric viruses transmitted via fecal-oral routes. Despite the substantial health impacts of NoV and EV, tracking regional and national infection dynamics remains challenging. Current surveillance systems focus on case clusters and severe hospitalized infections^1,2^, which likely represent only a fraction of the overall incidence in the community. Here we use wastewater-based surveillance (WBS) to complement clinical reports and demonstrate effective community-level surveillance of these enteric pathogens during the Coronavirus disease 2019 (COVID-19) pandemic in Switzerland.

NoV is one of the leading causative agents of gastroenteritis outbreaks in all ages, accounting for 10-20% of all acute gastroenteritis cases globally^3–5^. Among the human NoV genogroups, genogroup II (GII) is currently the most prevalent in clinical cases^6^, followed by genogroup I (GI)^7,8^. Human non-polio EV causes a broad spectrum of illnesses, including gastroenteritis, meningitis, acute flaccid myelitis, myocarditis and hand-foot-mouth disease. Young children are especially affected and exhibit the highest morbidity rates. Approximately 15,000 EV infections were reported between 2015 to 2017 across the EU and EEA^9^, though the true number of EV infections is likely higher. In the United States, an estimated 10-15 million EV infections occur annually^10^.

Wastewater-based surveillance monitors focal pathogens in municipal wastewater, typically using molecular methods characterized by high target specificity. It allows for passive and comprehensive monitoring of the entire population within a catchment area, regardless of symptom presentation or clinical testing access. Pioneered for enteric viruses, including NoV, poliovirus, and other enteroviruses^11–14^, this method gained prominence and widespread adoption during the COVID-19 pandemic^15,16^. Multiple studies have demonstrated strong spatial and temporal correlation between wastewater viral concentrations and clinical cases for a variety of pathogens^17–20^.

Wastewater-measured pathogen levels are also informative of important epidemiological parameters, such as the effective reproductive number, Re^21–24^. Wastewater-based Re values correspond well to those estimated based on clinical data when this is available^21^, and extend the ability to estimate Re to non-notifiable diseases with scarce clinical data^22^. To date, estimates of Re from wastewater have been restricted to respiratory pathogens. However, we demonstrate that the approach is readily extensible to common, non-notifiable enteric pathogens.

Beyond surveillance, longitudinal Re estimates are useful to study how external factors, like non-pharmaceutical interventions (NPIs), affect pathogen transmission dynamics. During the COVID-19 pandemic, school closures and movement restrictions reduced transmission of the respiratory pathogen SARS-CoV-2^25,26^ (the target of the interventions), as well as other respiratory viruses including influenza virus^27–29^ and respiratory syncytial virus^30^. Studies suggest the incidence of enteric pathogen infections (including NoV and EV) was lowered as well^31–34^. However, the efficacy of NPIs against enteric virus transmission remains underexplored and qualitative. Here we used our wastewater-based Re estimates for NoV and EV during the COVID-19 pandemic to evaluate the effect of NPIs on these enteric pathogens. This enhances our understanding of the effect of the COVID-19 pandemic on the transmission of other viruses, and may inform future interventions to mitigate the spread of NoV and non-polio EV.

## Methods

### Sample collection and storage

A total of 372 daily 24-h flow-proportional influent samples (after fine screening) were collected between December 5, 2020 and October 10, 2022 from Zurich’s wastewater treatment plant (WWTP, Werdhölzli) serving approximately 471,000 people. After collection, the wastewater samples were shipped on ice and stored at 4 °C for up to 8 d before processing.

### Sample concentration and RNA extraction

The samples collected before November 10, 2021 were processed by ultrafiltration followed by RNA extraction using QIAamp viral RNA Mini Kit (protocol 1). Samples collected after November 30, 2021 were processed by direct capture nucleic acid extraction (protocol 2). Samples processed between November 10 and November 30, 2021 were subjected to both protocols to compare the recovery efficiency of each. The extracted RNA was stored at -80°C for up to 1.5 yrs.

#### Ultrafiltration (Protocol 1)

Aliquots of 50-70 ml were spiked with murine hepatitis virus as a processing control. The mixed samples were clarified by filtration (2 µm glass fiber filter (Millipore) followed by a 0.22 µm filter (Millipore) for samples prior to March 8th and by centrifugation (30 min at 4200 x g) for samples after. Clarified samples were concentrated using centrifugal filter units (Centricon Plus-70 Ultrafilter, 10 kDa, Millipore) by centrifugation at 3,000 × g for 30 min. Centricon cups were inverted and the concentrate was collected by centrifugation at 1,000 × g for 3 min. The resulting concentrate (up to 280 µl) was extracted using the QiaAmp viral RNA mini kit (Qiagen) according to the manufacturer’s instructions and as previously reported^21^, adapted to the larger volumes, and eluted in 80 µl.

#### Direct Nucleic Acid Capture (Protocol 2)

Aliquots of 40 ml were spiked with murine hepatitis virus as a processing control. The mixed samples were lysed and eluted in 80 µl using the Wizard Enviro TNA Kit (Promega), according to the manufacturer’s instructions, and further purified using a OneStep PCR Inhibitor Removal Kit (Zymo Research, Irvine, CA, USA) as previously reported^22^.

### One-step reverse transcription-digital PCR (RT-dPCR)

The nucleic acid extracts were analyzed for SARS-CoV-2, NoV and EV, whereby SARS-CoV-2 was measured in daily extracts and NoV and EV every other day. SARS-CoV-2 RNA was quantified by RT-dPCR using the Naica System (Stilla Technologies, France) targeting the N gene marker N1^21^.

NoV and EV were quantified by duplex RT-dPCR assay using QIAcuity One (Qiagen, USA) as detailed by Li et al.^35^. The EV assay uses a reverse primer from a region conserved in human enteroviruses but absent in sequenced rhinoviruses and bovine enteroviruses, allowing for specific detection of human EV^36^. In each dPCR run, a negative control (no template control) and a positive control were included. The positive control was synthetic DNA (gBlock, Integrated DNA Technologies, USA) containing the target sequences. Quantities were expressed as genome copies (gc/µL) per reaction. The RT-dPCR assays were performed using automatic settings for the threshold and baseline for the EV and NoV concentrations, and manual gating for the SARS-CoV-2 concentrations. Samples with >2 positive partitions in both replicate wells and <5-fold difference between the duplicates were considered valid. A total of 363 out of 372 samples met these criteria and were subjected to further analysis. To transform the measured genome copies per reaction into absolute viral loads for the catchment, we accounted for sample volume processed (40-70 ml), elution volume (80 μl), eluate dilution (1:10), reaction template volume (4 μl), and influent flow rate of the WWTP (varied daily, in L)^21,22^.

### Obtaining epidemiological parameters from the literature

We reviewed the literature to obtain estimates for epidemiological distributions that are important to describe viral transmission dynamics (Table 1, Figure S1). To derive effective reproductive number (Re) estimates from wastewater, this includes the shedding load distribution (SLD) and the generation time interval distribution^21^. The SLD describes the probability that an average infected individual sheds a given virus particle, *t* days after infection. The generation time interval distribution describes the distribution of observed time intervals between the infection of an individual *j* and the infection of its infector *i*.

**Table 1:**
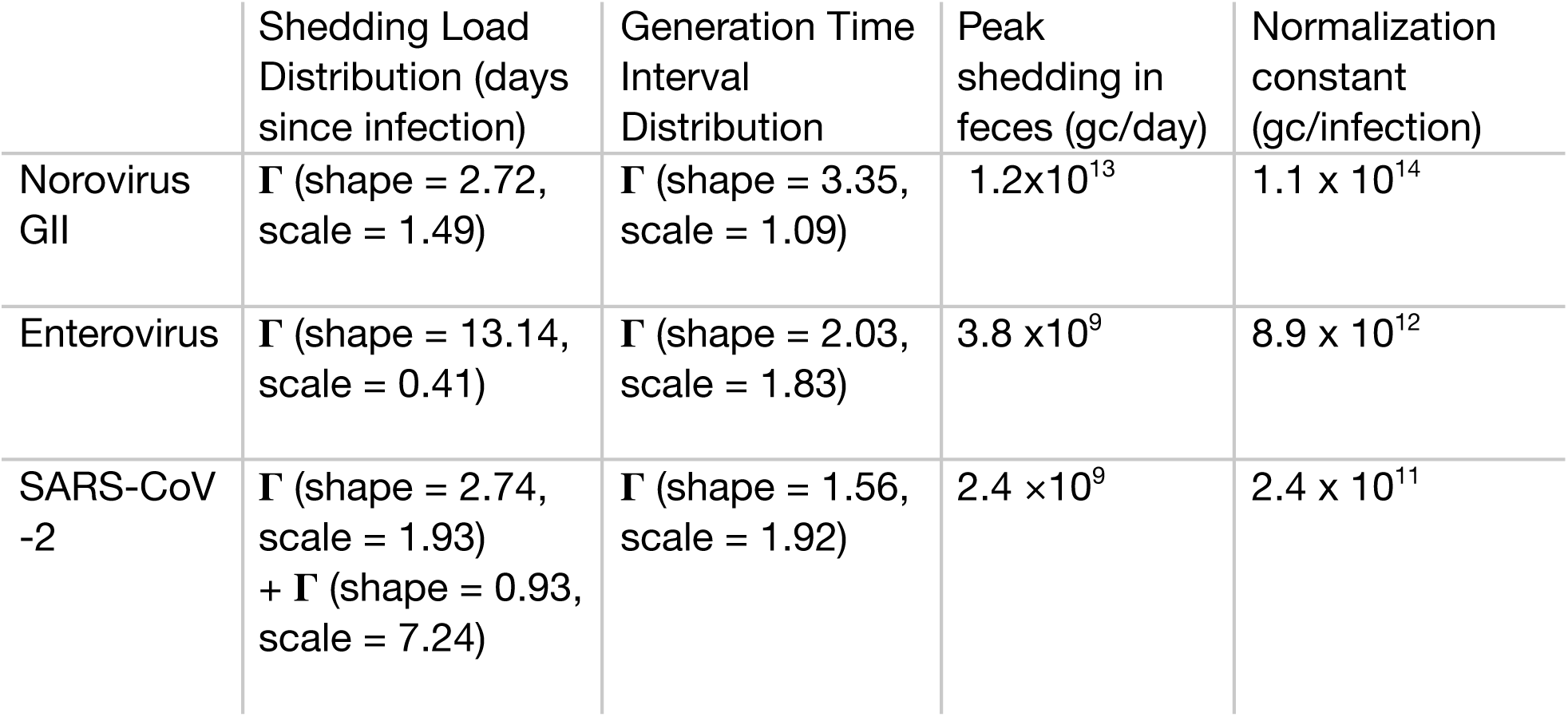
Assumed shedding load distributions, generation time intervals, peak shedding in feces (gc/day), and normalization constants (gc/infection) for the estimation of infection incidence and effective reproductive number (R_e_) for NoV, EV, and SARS-CoV-2. The shedding load distribution for SARS-CoV-2 consists of two components: a distribution from infection to symptom onset, and from symptom onset to shedding. Distributions are visualized in Supplementary Figure S1. The normalization constants (gene copies per infection) correspond to the lowest measured viral loads for each virus during the observation period.

To estimate the SLD for NoV GII, we used data from a human vaccine trial^37^. Healthy adults were challenged with GII.4 virus and screened for fecal shedding over the following ten days. We fitted a gamma distribution (with mean 4 days, standard deviation, sd, 2.5 days) to shedding data from the placebo arm of the study which contained participants that were not vaccinated (data digitized from Fig. 2 in Bernstein et al^37^). The amount of virus shed upon infection was taken from a similar trial with controlled human infection: median peak shedding was found to be 95×10^9^ gene copies (gc) per gram feces (range 0.5-1640 ×10^9^ gc/g)^38^. Given the average amount of feces per person per day (128g, range 51–796g^39^), this yields a lower bound of 1.2×10^13^ (range 2.6×10^10^ - 1.3×10^15^) gc/infection. The generation time interval was estimated from data of NoV transmission in day care centers in Sweden^40^, analogous to Heijne *et al.*^41^. We estimate a gamma distribution with a mean of 3.1 days, and sd 1.8 days. This is slightly longer than the mean serial interval, the time between symptom onset of two people in a transmission chain, of 2.2 days observed in the same outbreak^40^, or of 1.86 days (95% confidence intervals: [1.6, 2.2]) estimated during hospital outbreaks in England^42^. The median incubation period, the time from exposure until symptom onset, is also shorter, and has been estimated at 1.5 days in Sweden^40^ or during an outbreak among American football players^43^. A 2013 systematic review found that the mean incubation period of both NoV GI and GII is 1.2 days^44^.

To estimate the SLD for EV, we used data from an enterovirus 71-coxsackievirus A16 (EV71-CA16) vaccine study in rhesus macaques^45^. We fitted a gamma distribution (mean 5.4 days, sd 1.5 days) to viral titers in oral swabs from the placebo arm of the study (data digitized from Fig. 3B of Fan et al.^45^). We used EV71-CA16 as a stand-in for other EVs, because they were the only non-polio enteroviruses for which we could find data to estimate the SLD. As an approximation of the generation time interval distribution, we used the serial intervals observed in EV71 transmission between household contacts in Taiwan^46^. The authors reported only the mean of 3.7 days and sd of 2.6 days of the serial interval distribution, rather than its functional form. We assume a gamma distribution for compatibility with the estimateR method and analogy with the other distributions used in this study (previous studies have assumed a Weibull distribution based on these data^47,48^). We note that enteroviruses other than EV71 may have notably different SLDs or serial intervals.

To estimate the number of EV gene copies shed during infection, we resort to data from tissue culture infection studies. We did not find reports of shedding loads measured by RT-PCR. Data suggests that throughout the infection non-polio EVs are shed at 2.0-5.5 log_10_ TCD_50_ (tissue culture dose) per gram feces^49^. However, the concentration of virus that would be picked up by RT-PCR may be 10-100 times higher^50^, i.e. 3.0-7.5 log_10_ gc/g feces. This is similar to the median viral loads of 4±9×10^6^ gc/g feces reported in infections with human rhinoviruses^51^, or 1.8×10^7^ gc/g feces reported for Aichi virus, a close relative of the enteroviruses^52^. Assuming that the wastewater signal will be dominated by the day of maximal shedding, this would suggest a lower bound of 3.8 ×10^9^ gc/infection.

For SARS-CoV-2 we follow the parameters used in Huisman et al^21^. For the SLD we use a combination of the incubation period by Linton et al (gamma distributed with mean= 5.3 days, sd = 3.2 days) and the shedding load distribution in days since symptom onset from Benefield et al. (gamma distributed with mean=6.7 days, sd= 7.0 days)^53,54^. For the generation time interval distribution, we use an estimate of the serial interval distribution from Nishiura et al (gamma distributed with mean= 4.8 days, sd = 2.3 days)^55^. In the time period under consideration, the Alpha, Delta, and Omicron variants circulated in Switzerland. Omicron BA.1 was associated with slightly lower viral shedding than Alpha, Delta, and Omicron BA.2^56^. Both Delta and Omicron BA.1 have shorter incubation periods than ancestral SARS-CoV-2 (3.7-4 days for Delta and 3-3.4 days for Omicron respectively, compared to 4.6-6.4 days for the ancestral strain^56^). However, in this study, we do not assume any differences in SLD or generation time interval distribution due to the appearance of variants. We assume a population-average shedding load distribution and do not take into account the effect of superspreaders, disease severity, or vaccination status on the amount or dynamics of viral shedding^56^. For SARS-CoV-2 the median peak shedding value found was 1.9×10^7^ gc/g feces^57^; corresponding to at least 2.4 ×10^9^ gc/infection.

### Clinical data of tests for gastrointestinal viruses

Noro- and enteroviruses are not notifiable diseases in Switzerland, and therefore there is no centralized surveillance system for these etiologies. However, some large hospitals in Switzerland screen patients with gastrointestinal symptoms for the presence of major causative agents of gastrointestinal disease, including EV and NoV GII. These tests are conducted for individual diagnostics based on the prescription of a physician. We obtained such epidemiological data from the Hôpitaux Universitaires Genève (Geneva University Hospitals, HUG)^58^, the largest hospital in Switzerland, on RT-PCR gastrointestinal virus panels performed over the past 12 years (from Jan 2012 to May 2024). We further obtained data on all tests conducted to screen for noro- and enteroviruses at the Institute of Medical Virology, University of Zurich (IMV/UZH), between 23-04-2018 and 01-11-2022. This lab conducts tests for both the University Hospital Zurich, the largest hospital in our catchment area, and external partners. Tests were performed on GeneXpert, Biofire and Ridagene gastrointestinal panels and with in-house RT-PCR. In this case, commercial RT-PCR tests could not distinguish between entero- and rhinoviruses, so the reported numbers correspond to the total of both etiologies. Both HUG and IMV/UZH datasets contain all negative, positive, and non-assessable PCR-tests analysed per day. Testing data may overrepresent outbreak clusters, in particular involving nosocomial transmission.

### Estimating the infection incidence and effective reproductive number

In our primary analysis, we use the estimateR package (v0.2.1) in R to estimate the infection incidence and the effective reproductive number from the longitudinal wastewater measurements^59^, analogous to Huisman et al.^21^

First, we interpolated missing values and smoothed the measured viral loads for all three viruses with a 28-day sliding window. To obtain values in the right order of magnitude, all measured loads were normalized by a normalization constant defined as the lowest viral load measured for the respective virus (Table 1). Next, we used estimateR to deconvolve the smoothed viral loads by the virus-specific shedding load distribution (Table 1) and obtain the estimated infection incidence (Supplementary Fig S2). In a second step, the software uses EpiEstim to estimate the effective reproductive number from the infection incidence^60^. In this step we used the virus specific generation time interval distributions listed in Table 1.

For sensitivity analysis of the inference pipeline, we further estimated R_e_ using the EpiSewer package^61^ (v0.0.3), a Bayesian generative modeling framework to estimate Re from wastewater loads. This package is still in beta testing, but proves complementary as it assumes a different noise model for wastewater viral load observations, leading to different uncertainty interval estimates, and explicitly models the effect of a limit of detection during dPCR, which affects the estimates during periods of low infection incidence. For the estimates with EpiSewer, we assumed the same parameters as for the estimateR pipeline, including shedding load and generation time interval distributions (Table 1), and viral load per case used as normalization constants (Table 1).

### Estimating the effect of non-pharmaceutical interventions

To estimate the effect of non-pharmaceutical interventions on the viral transmission dynamics, we use the KOF stringency index^62^. This index records when Swiss authorities introduced and lifted public health interventions to limit the spread of SARS-CoV-2 in Switzerland (Table S1). The index is based on information from nine sub-indicators that include school, workplace or transport closure, restrictions on gatherings and public events, stay-at-home requirements, restrictions on within-country or international travel, and public information campaigns. We define days on which the KOF stringency index changed as “changepoints”. To assess the effect of a change in stringency on disease transmission, we computed the change in the effective reproductive number Δ*R*_e_ = *R*_e_ (*t*) − *R*_e_ (*t* + 14) for each virus in the 14 days after a changepoint.

### Data availability

All data and code to reproduce the figures are available on github: https://github.com/JSHuisman/noro_entero_wastewater. This repository also includes a vignette with more general instructions on how to estimate R_e_ from wastewater viral loads.

## Results

### Sampling and determination of wastewater viral loads

We retroactively monitored NoV GII and EV in wastewater collected between 5 December 2020 and 10 October 2022 (Fig. 1A). The measured viral loads display clear peaks of viral shedding: NoV peaked in March 2021, September 2021, and November/December 2021. EV similarly peaked in March 2021, as well as August 2021 and July 2022. Over the same timespan, SARS-CoV-2 exhibited minor peaks in April and August 2021 and January 2022, and two large peaks in March and July 2022.

**Figure 1:**
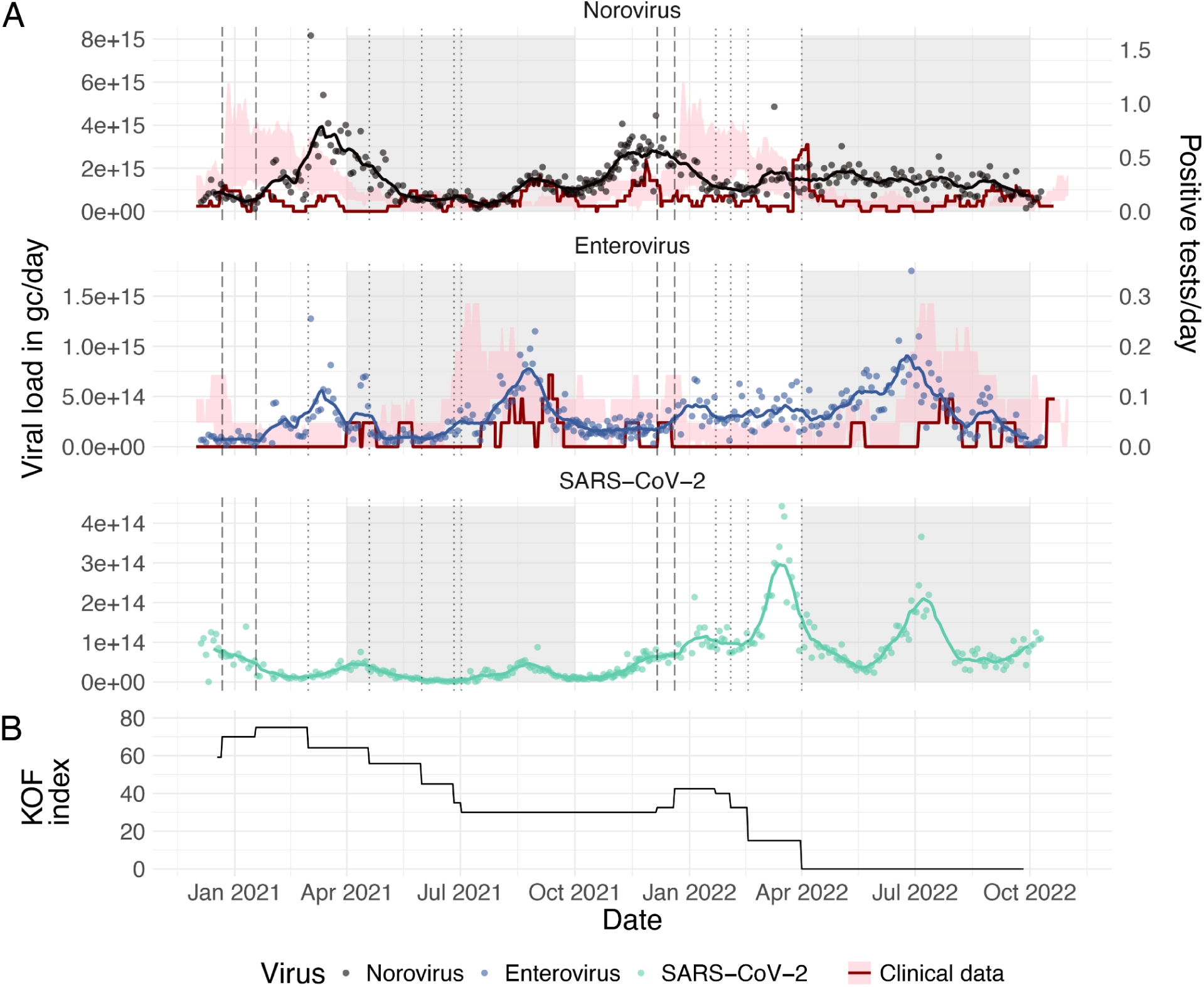
**A)** Measured viral loads for Norovirus GII (top, black points), Enterovirus (middle, blue points) and SARS-CoV-2 (bottom, green points) in wastewater collected at the Werdhölzli wastewater treatment plant (Zurich). The continuous lines are 21-day smoothed averages of the measured viral loads. Pink ribbons indicate the historical average number of positive tests for each virus at the Geneva university hospitals (25th, 75th quantile) across the years 2012-2019, 2023, 2024. The red line indicates the number of positive tests observed at the IMV/UZH during our study. Clinical cases were averaged over a 21-day sliding window (right-hand axis). Grey shaded regions indicate the spring and summer months (Apr-Sep), white regions indicate fall and winter months (Oct-Mar). Dashed vertical lines indicate increases in COVID-19 containment measures, and dotted lines reductions in stringency. **B)** KOF stringency index (0-100) indicating the strength of non-pharmaceutical interventions in place.

Increases in the wastewater signal of NoV largely coincided with an increase in the number of clinically observed NoV cases (Fig 1A, red line). Peaks observed in clinical cases always coincided with elevated viral loads in the wastewater. This was most clear in September and November/December 2021. Notably, the signal from clinically observed cases was more variable than in wastewater, with frequent periods in which no cases were detected. The relative magnitude of peaks also differed between the wastewater and clinical data.

NoV loads were very low in January 2021 with a peak only occurring in March 2021. This timing differs from historical trends, which shows a marked peak in clinical observations in the winter months (Fig 1A, pink 10-yr average; Fig S2, S3). While clinical observations remained low until September 2021, the wastewater data shows clear evidence of transmission in March that year. NoV transmission thus seems to have been delayed compared to its usual (winter) seasonality. In 2022, the peak preceded the typical timing of NoV transmission by several months.

Peak EV loads in wastewater largely followed the summer seasonality expected based on the long-term clinical average (Fig 1A, S2, S3), with a notable additional peak in EV loads observed in March 2021. Since our dPCR assay targets a conserved region of the EV genome (the 5’-non coding region), the observed wastewater loads are the sum total of loads of human EVs. The observed peaks may thus describe outbreaks caused by different *Enterovirus* genotypes. Interestingly, the wastewater EV peaks that were accompanied by clinical cases (Apr 2021, Sep 2021 and Jul 2022) largely coincided with SARS-CoV-2 peaks. This may be explained by co-infections with both viruses^63,64^. The peak in Sep 2021 coincided with a Europe-wide wave of clinically reported EV infections between Sep-Nov 2021^65^. The European Non-Polio Enterovirus Network (ENPEN) reported that 9.6% of these infections were due to EV-D68, which is notable due to its association with acute flaccid myelitis and severe respiratory outcomes. However, of 74 enterovirus-positive samples sequenced at IMV/UZH between July to December 2021, only 1 was ascribed to EV-D68^66^.

### Estimating the infection incidence and effective reproductive number

To better assess the dynamics of enteric virus transmission, we estimated the effective reproductive number, R_e_, from the measured wastewater viral loads (Fig 2). The observed peaks in wastewater viral loads correspond to periods during which R_e_ > 1. NoV and EV both reached maximum values of R_e_ within one week after an increase in non-pharmaceutical interventions on January 18 2021. The maximum value attained was R_e_ = 1.28 (0.86, 1.78) for NoV and R_e_ = 1.42 (1.04, 1.84) for EV respectively. During the rest of our observation period, both genera attained maximum R_e_ values between 1.1-1.2, with slightly higher R_e_ estimates for EV (Fig 2; Table 2). EV also showed more variable transmission dynamics than NoV. Averaged over longer periods of time both genera displayed an R_e_ of 1, as would be expected for endemic viruses. NoV reached quarterly average values above 1 in Q1 2021 and Q3/Q4 2021. EV reached quarterly averages above 1 in Q1/2/4 2021 and Q2 2022. Notably, R_e_ ≥ 1.2 for EV were attained in Q1/Q2/Q3 2021 and Q3 2022. These trends were also observed using the modeling framework from the EpiSewer package, suggesting they are independent of the method used to estimate R_e_ (Fig. S6, Table S2).

**Figure 2:**
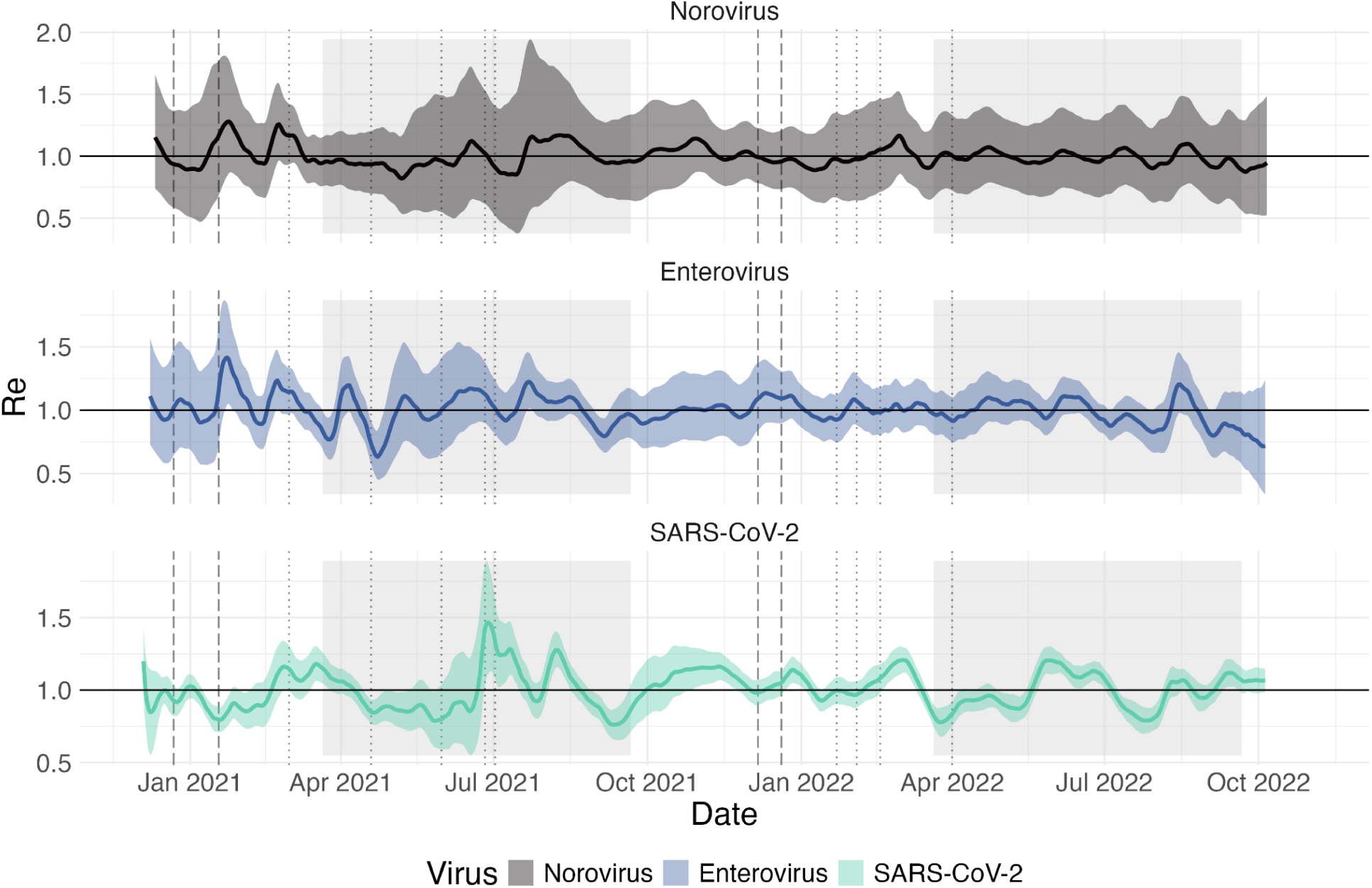
Estimated effective reproductive number (Re) for NoV (top, black), EV (middle, blue), and SARS-CoV-2 (bottom, green) using the estimateR pipeline. Grey shaded regions indicate the spring and summer months, white regions indicate fall and winter months. Dashed vertical lines indicate increases in the KOF stringency index, which is an indicator of COVID-19 containment measures, and dotted lines indicate reductions in stringency. Wastewater viral load data were normalized and smoothed over a 28-day window prior to Re estimation.

**Table 2:**
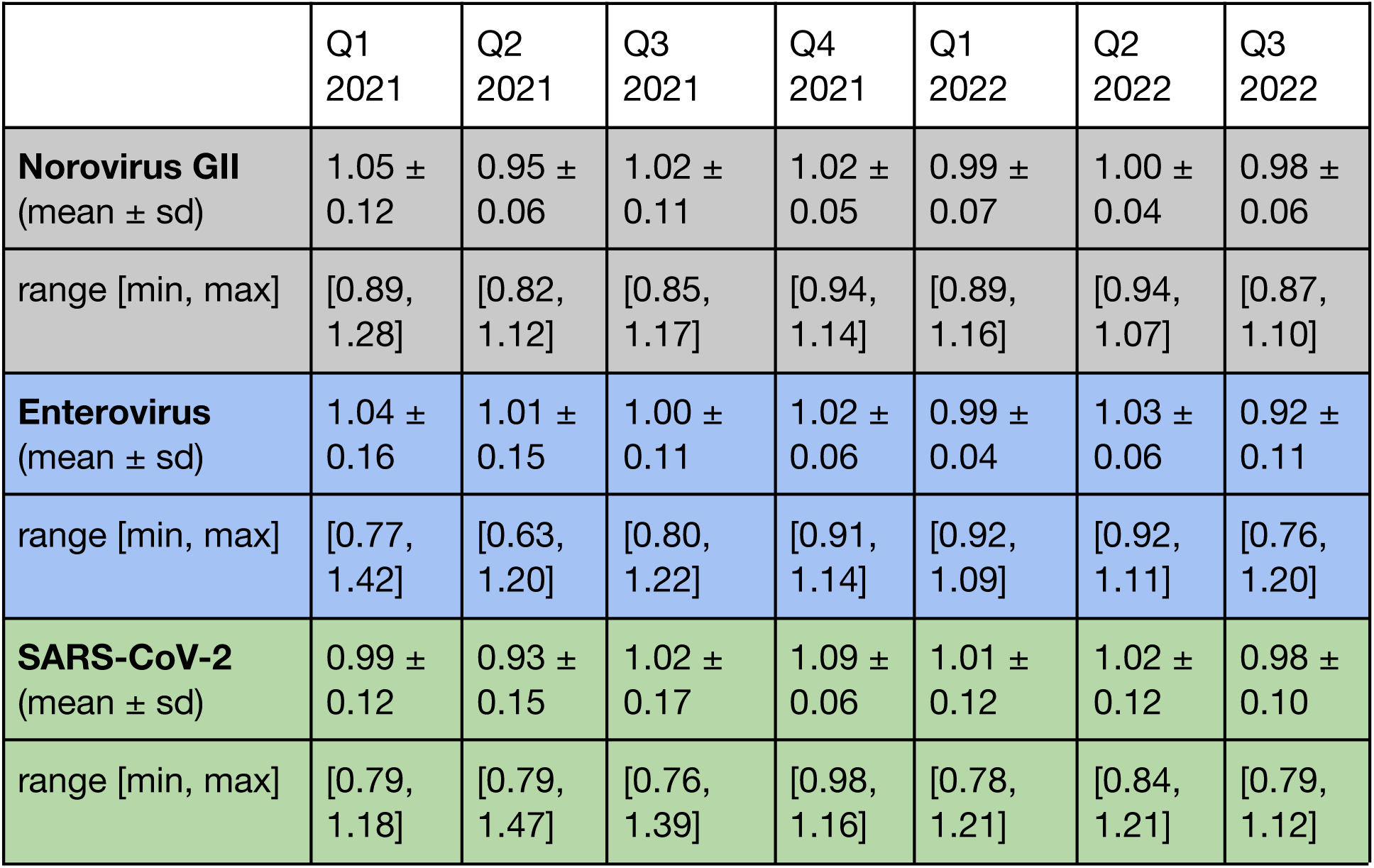
Quarterly estimates of the effective reproductive number of Norovirus GII, Enterovirus, and SARS-CoV-2. For each virus, the first row corresponds to the average R_e_ per quarter, and the second row to the range of values observed during that time window.

|  | Q1<br>2021 | Q2<br>2021 | Q3<br>2021 | Q4<br>2021 | Q1<br>2022 | Q2<br>2022 | Q3<br>2022 |
| --- | --- | --- | --- | --- | --- | --- | --- |
| **Norovirus GII**<br>(mean ± sd) | 1.05 ±<br>0.12 | 0.95 ±<br>0.06 | 1.02 ±<br>0.11 | 1.02 ±<br>0.05 | 0.99 ±<br>0.07 | 1.00 ±<br>0.04 | 0.98 ±<br>0.06 |
| range [min, max] | [0.89,<br>1.28] | [0.82,<br>1.12] | [0.85,<br>1.17] | [0.94,<br>1.14] | [0.89,<br>1.16] | [0.94,<br>1.07] | [0.87,<br>1.10] |
| **Enterovirus**<br>(mean ± sd) | 1.04 ±<br>0.16 | 1.01 ±<br>0.15 | 1.00 ±<br>0.11 | 1.02 ±<br>0.06 | 0.99 ±<br>0.04 | 1.03 ±<br>0.06 | 0.92 ±<br>0.11 |
| range [min, max] | [0.77,<br>1.42] | [0.63,<br>1.20] | [0.80,<br>1.22] | [0.91,<br>1.14] | [0.92,<br>1.09] | [0.92,<br>1.11] | [0.76,<br>1.20] |
| **SARS-CoV-2**<br>(mean ± sd) | 0.99 ±<br>0.12 | 0.93 ±<br>0.15 | 1.02 ±<br>0.17 | 1.09 ±<br>0.06 | 1.01 ±<br>0.12 | 1.02 ±<br>0.12 | 0.98 ±<br>0.10 |
| range [min, max] | [0.79,<br>1.18] | [0.79,<br>1.47] | [0.76,<br>1.39] | [0.98,<br>1.16] | [0.78,<br>1.21] | [0.84,<br>1.21] | [0.79,<br>1.12] |

The R_e_ of NoV and EV were correlated throughout our observation period, though neither of the enteric viruses showed a significant correlation with the transmission dynamics of SARS-CoV-2 (Fig. S7). Here we averaged the R_e_ for each week in our observation period to reduce the bias stemming from autocorrelation between R_e_ on different days. The observed correlation between EV and NoV is surprising since NoV is normally associated with winter transmission whereas EV is typically associated with summer transmission. We speculate that the observed correlation stems from a similar impact of non-pharmaceutical interventions and other disruptions during the COVID-19 pandemic on virus transmission.

### The effect of non-pharmaceutical interventions on enteric virus transmission

The period of our study was marked by substantial non-pharmaceutical interventions to curb the spread of SARS-CoV-2 (Fig 1B). Using our measurements of NoV and EV wastewater viral loads and effective reproductive numbers, we can estimate the impact of these NPIs on enteric virus transmission and compare them to the impacts on SARS-CoV-2.

Both viral loads and Re estimates (Fig 1, 2) show that NoV transmission peaked later than usual in March 2021 and earlier than usual in September 2021 (Fig. S2, S3). In contrast, EV transmission largely followed the expected summer seasonality, apart from anomalous transmission in Jan-March 2021. This suggests that the restrictions on public events and workplace closings in Jan 2021 delayed transmission of both viruses.

To investigate the impact of implementing or lifting individual NPIs, we analyze how a change in the KOF stringency index (ΔKOF) affects the Re (ΔR_e_) 14 days later. In general, we observe increases in the Re for NoV and EV following decreases in NPI stringency and decreases in Re following increases in stringency (Fig. 3A, B). This pattern suggests that the NPIs reduce NoV and EV transmission. However, the observed increase in Re for SARS-CoV-2 following increases in stringency does not mean NPIs did not work for SARS-CoV-2. More sophisticated statistical analyses over different time periods and multiple geographic regions have shown that NPIs were effective at reducing SARS-CoV-2 transmission^25,62,67^. However, as the timing of the NPIs in Switzerland was tied to the epidemiological situation for SARS-CoV-2, but not NoV or EV, there may be biases in our SARS-CoV-2 analysis. For example, NPIs were generally implemented during times in which public awareness of SARS-CoV-2 case numbers was high, such that people may have changed their behaviour (and thus Re) already prior to the implementation of new measures. Unfortunately, the low number of independent observations of stringency changes and their corresponding effect on virus transmission prevent us from studying these trends in greater detail.

**Figure 3:**
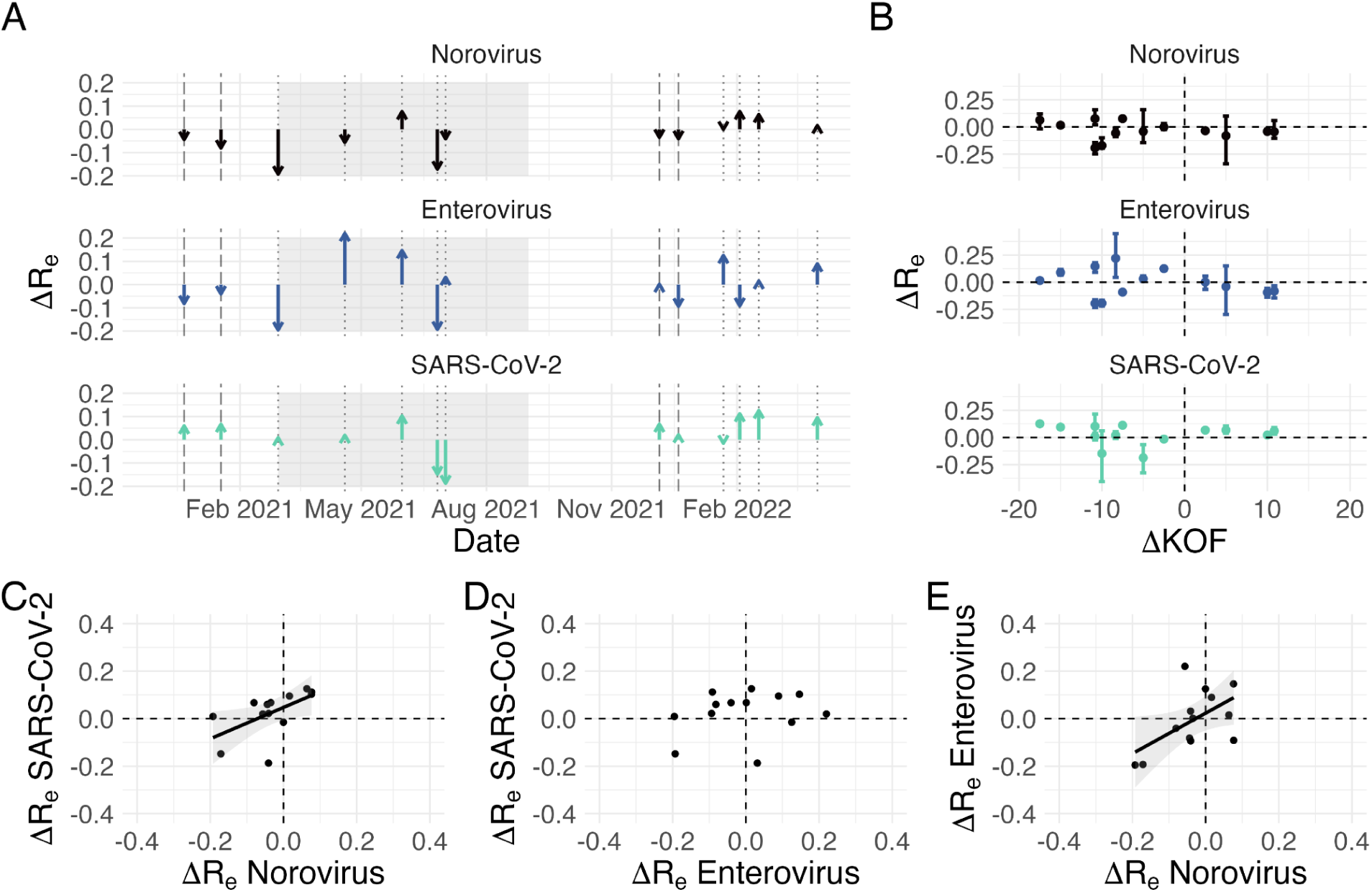
Change in Re (ΔR_e_) 14 days after a change in the KOF stringency index (ΔKOF). A) Dashed vertical lines indicate days with increasing measures, dotted lines indicate relaxation. Magnitude of the arrow indicates ΔR_e_ 14 days after that changepoint. B) The same ΔR_e_ values as in panel A, but shown as a function of the change in stringency at the corresponding changepoint (14 days prior). The error bars indicate the change in the upper and lower 95% confidence interval over 14 days. C-E) Correlation between ΔR_e_ for the different viruses.

Given the correlation between the R_e_ estimates of NoV and EV throughout our observation period (Fig. S7), it is not surprising that the change in Re after a KOF changepoint is also significantly correlated between NoV and EV (Fig. 3E). At these specific timepoints, the ΔR_e_ of NoV and SARS-CoV-2 is also correlated, while the correlation between EV and SARS-CoV-2 was not significant (Fig. 3C, D).

## Discussion

We demonstrate that measuring wastewater viral loads of NoV and EV provides valuable insights for pathogen surveillance and real-time monitoring of transmission dynamics. Wastewater did not only recapitulate enteric virus transmission that was reflected in an uptick in clinical cases, but also demonstrated transmission that went previously undetected. The wastewater data further proved useful to estimate the population-level effective reproductive number of both NoV and EV, and to assess the impact of non-pharmaceutical interventions on the transmission of these enteric viruses during the COVID-19 pandemic in Switzerland.

Since wastewater provides a continuous source of population level information on disease transmission, it extends existing knowledge about the dynamics of enteric viruses. Previous sources of information stem primarily from hospitals and outpatient care. A study in Switzerland found that these sources underestimate the burden of acute gastroenteritis^68^. Clinical testing was preferentially prescribed for patients with a prolonged duration of illness, general ill health, recent travel abroad, or during ongoing outbreaks. This may explain why the wastewater and clinical data showed a different relative magnitude of transmission peaks. Another explanation could be shifts in the demographics of infection. For example, both health risk (reflected in clinical cases) and virus shedding (reflected in wastewater) could differ across age and sex. It is notable that our data showed a peak in both NoV and EV wastewater viral loads after release of COVID restrictions in Mar 2021, which was not reflected in the clinically observed cases.

Estimating Re for NoV and EV from wastewater also opens up the ability to understand transmission at the population, rather than the outbreak level. Outbreak data often centers on superspreading events which may lead to an overestimated Re. This could explain why our estimates of the maximum Re of NoV (around 1.1-1.2) are lower than values found for NoV in previous studies applying transmission models to outbreak data^69,70^. Our result for EV (Re = 1.1-1.2) is also on the lower end of the uncertainty interval reported for EV in the US^71^, yet close to the value estimated for hand-foot-and-mouth disease in Hong Kong (Re = 1.1–1.2)^47^.

Our Re estimates further suggested that EV follows more stochastic transmission dynamics than NoV. This is somewhat surprising since any viral transmission dynamics driven by superspreading events would lead to more stochasticity in R_e_ ^72^, as would low overall numbers of infections. Both would suggest that NoV dynamics should be more variable than EV. Our estimates of the infection incidence (Supp Fig. S5) are 10x higher for EV than NoV. This is likely even an underestimate of EV incidence, as our normalization constant is roughly 10^3^ times the peak viral load shed by a single individual (Table 1). However, the stochasticity of EV wastewater viral loads may be higher if the variation in shedding between people is high. This variability could for instance stem from differences in age, sex, immune or coinfection status. Unfortunately there is little data to support or discredit this hypothesis. Lastly, the stochasticity may be higher for EV because our assay provides an average R_e_ for all enteroviruses. Distinct EV genotypes may cause multiple overlapping waves leading to a more stochastic signal, in contrast to the higher specificity of the NoV assay targeting only genotype II. Future work should focus on estimating genotype-specific EV viral loads from wastewater to obtain Re estimates for individual EV genotypes.

Lastly, our Re estimates allowed us to investigate the effect of non-pharmaceutical interventions during the COVID-19 pandemic on the transmission dynamics of NoV and EV. We observed surprisingly low wastewater viral loads in winter 2021, when strong NPIs were in effect (Fig. 1,2). Low enteric virus prevalence during winter 2021 was also observed in clinical data in Austria^31^ and Australia, outbreak data in the US^34^, and in wastewater data from Austria and California^32^. Together with earlier than normal peaks of NoV transmission in Sep and Nov/Dec 2021, this led to correlated dynamics of EV and NoV during our study (Supp Fig S7). Given the well-known anti-correlation between EV and NoV, this suggests a strong disruption of normal seasonal patterns due to COVID-19 and related NPIs. Since NoV transmission is characterized by a relatively high proportion of superspreading events (especially when vomiting took place), NPIs that reduce the size of large gatherings and public events are especially likely to have a strong effect on transmission^41,69,72^. Unfortunately, our ability to estimate the impact of specific NPIs was limited by the generation of wastewater data from only one geographic location, with a total of 12 changes in NPI strength over the course of our observation period. In particular, we did not have wastewater data for early 2020, when the strongest interventions were in effect, and our observation period contains more decreases than increases in intervention stringency.

Wastewater measurements have great potential to improve understanding of the epidemiology of enteric viruses. Continued wastewater surveillance for pathogens is a powerful source of consistent, long-term infectious disease data, which provides an ideal testing ground to assess the impact of epidemiologically-relevant factors, such as non-pharmaceutical interventions, seasonality, or temperature on transmission. In addition, future work should focus on combining the wastewater signal with clinical and public health information on transmission clusters and clinical presentation, or social factors including school holidays, to uncover the factors that drive NoV and EV transmission at a population scale in the absence of pandemic disruption.

## Data Availability

Data and code to reproduce the figures are available on github: https://github.com/JSHuisman/noro_entero_wastewater.

## Acknowledgements

We gratefully acknowledge the efforts of Prof. Laurent Kaiser and Dr. Manuel Schibler in collecting the gastrointestinal virus data at the Geneva University Hospitals. We thank Dr. Pauline Vetter and Prof. Isabella Eckerle for facilitating access to this data. We further gratefully acknowledge Dr. Verena Kufner for her role in gastrointestinal virus screening at the IMV/UZH. We thank Franziska Böni, Alexander J Devaux, Xavier Fernandez-Cassi, Anina Kull, Christoph Ort, Elyse Stachler, and Blanche Wies for supporting the wastewater sample collection and processing. JSH was funded by the Human Frontier Science Program (HFSP) Postdoctoral Fellowship LT0045/2023-L. Further funding was provided by the Swiss National Science Foundation (Grant No. 196538), and a grant from the Swiss Federal Office of Public Health to TRJ and Christoph Ort.

## Supplementary Figures

**Figure S1:**
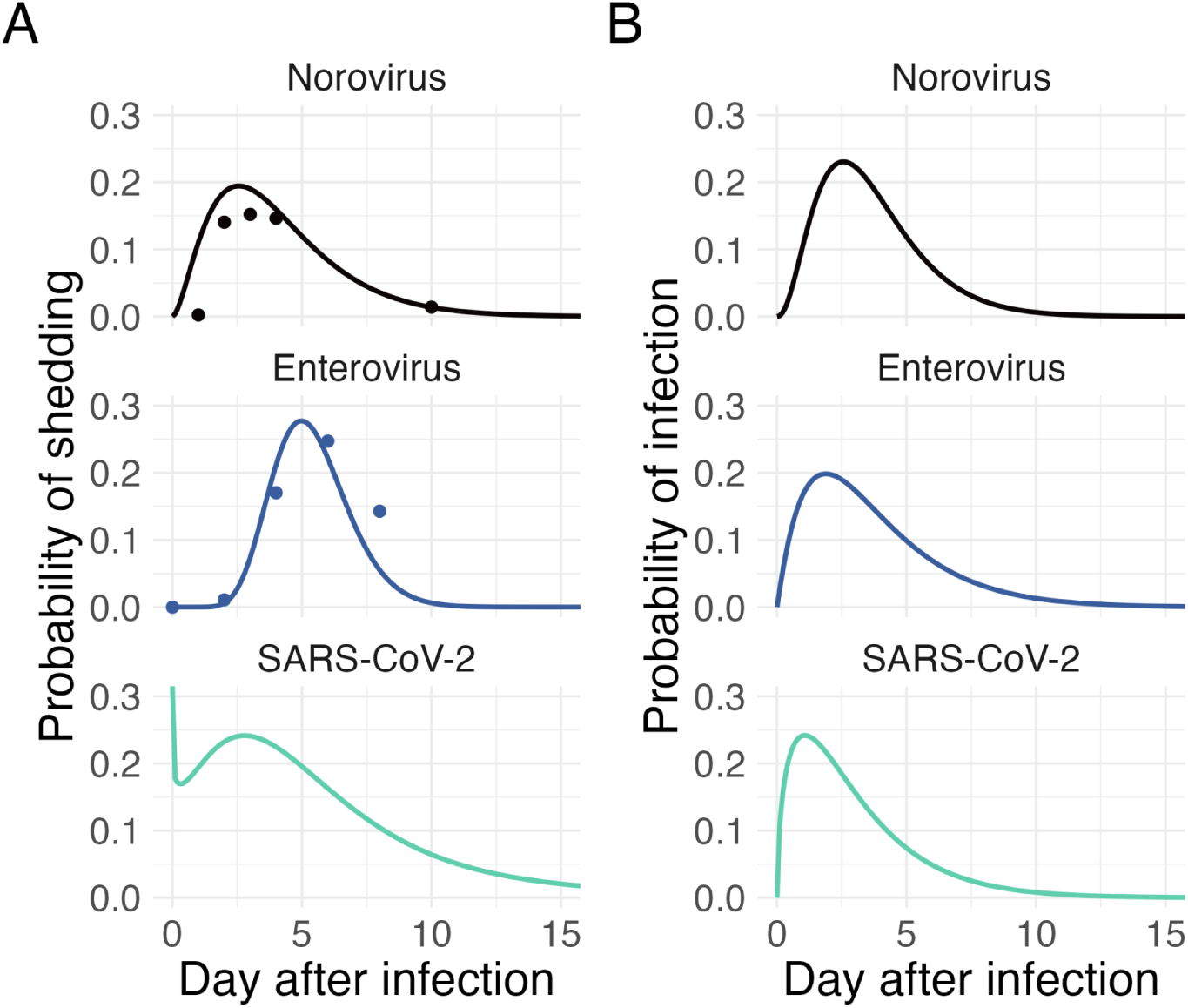
**A)** Assumed shedding load distribution for NoV (top, black; points represent data by Bernstein et al^37^), EV (middle, blue; points represent data by Fan et al^45^) and SARS-CoV-2 (bottom, green). **B)** Assumed generation time distribution for NoV (top, black), EV (middle, blue) and SARS-CoV-2 (bottom, green).

**Fig. S2.**
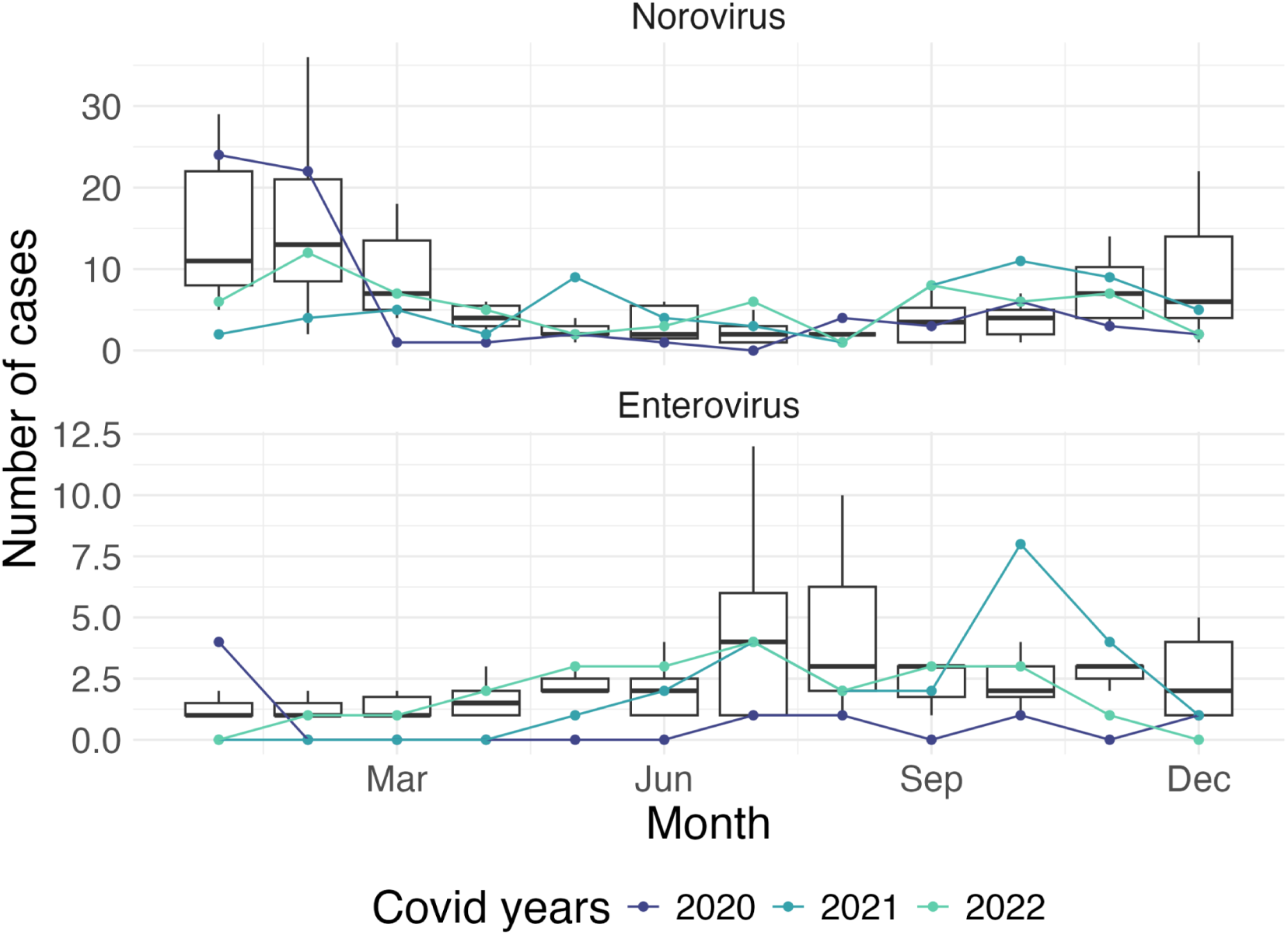
Yearly pattern of positive tests for NoV and EV at the Geneva University Hospitals. Boxplots summarize measurements made between 2012-2019 and 2023-2024. Colored lines indicate the number of positive tests during the COVID-19 pandemic (2020-2022).

**Fig. S3.**
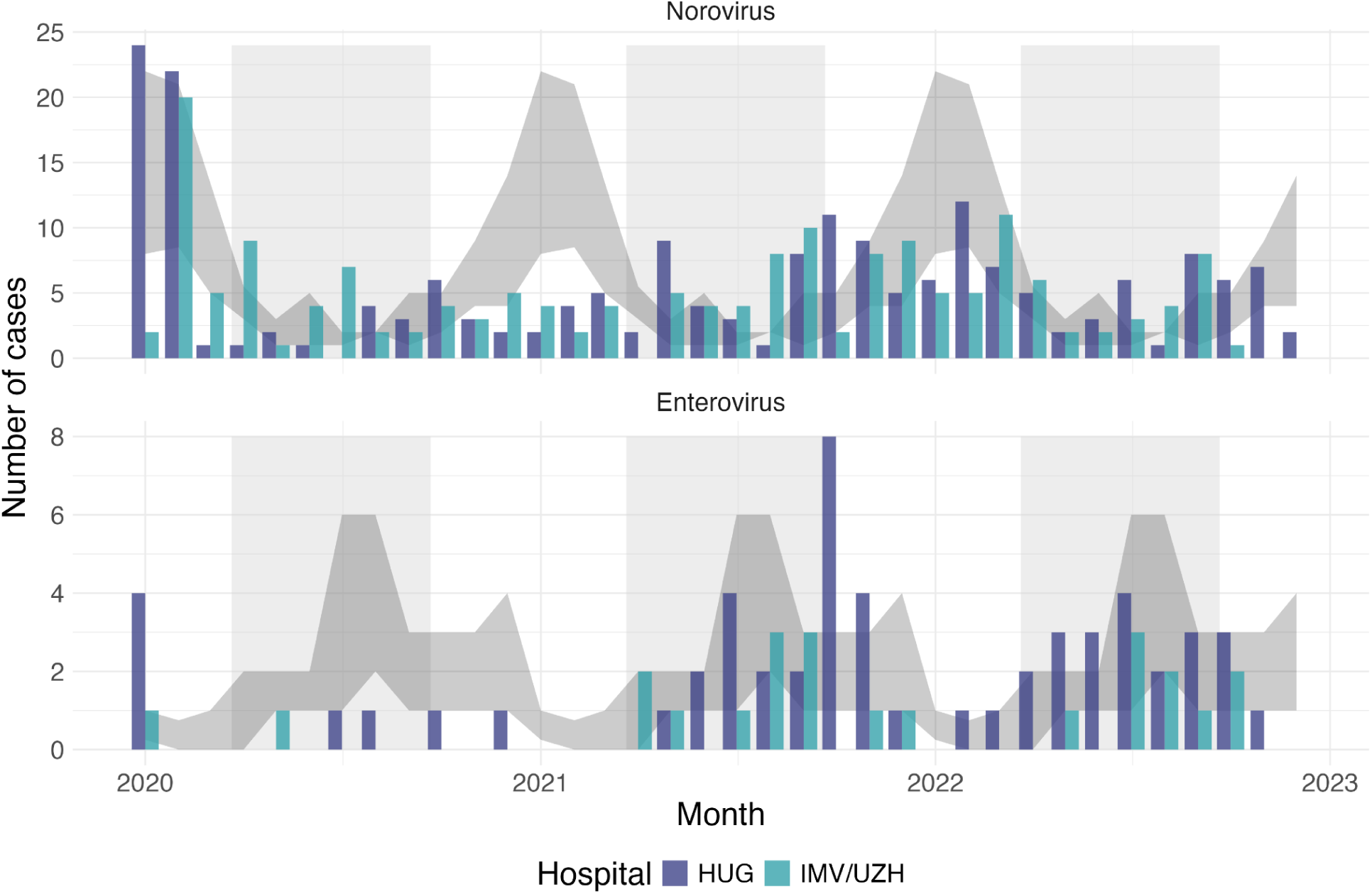
Number of positive tests for NoV and EV in the Geneva university hospitals (HUG, dark blue bars) and at the Institute for Medical Virology of the University of Zurich (IMV/UZH, turquoise bars). Cases are aggregated at a monthly level. The shaded regions indicate the average number of NoV or EV cases observed at HUG (25th, 75th quantile across years 2012-2019, 2023, 2024).

**Fig. S4.**
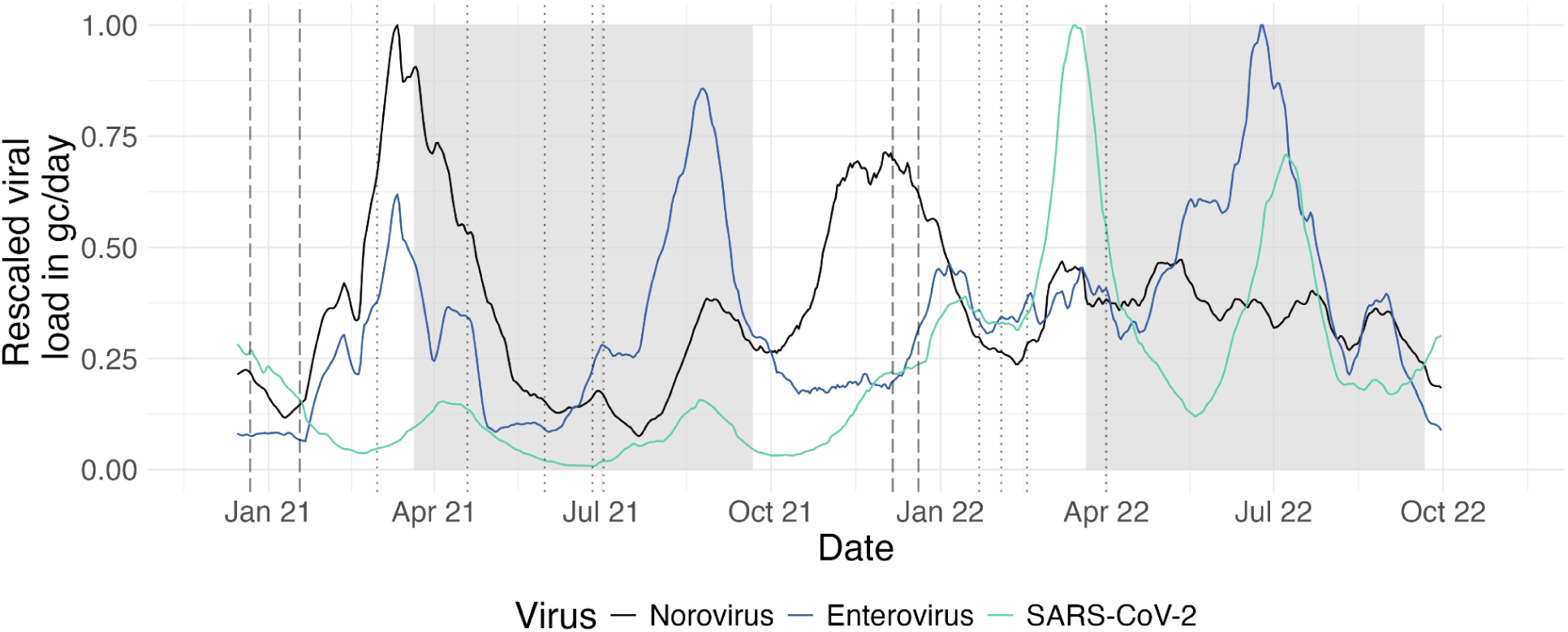
Rescaled viral loads (gc/day) for NoV, EV, and SARS-CoV-2 measured at the Zurich Werdhölzli wastewater treatment plant. Lines indicate the 21-day smoothed average of the measured viral loads. Values were rescaled by the maximal viral load observed for the respective virus.

**Figure S5:**
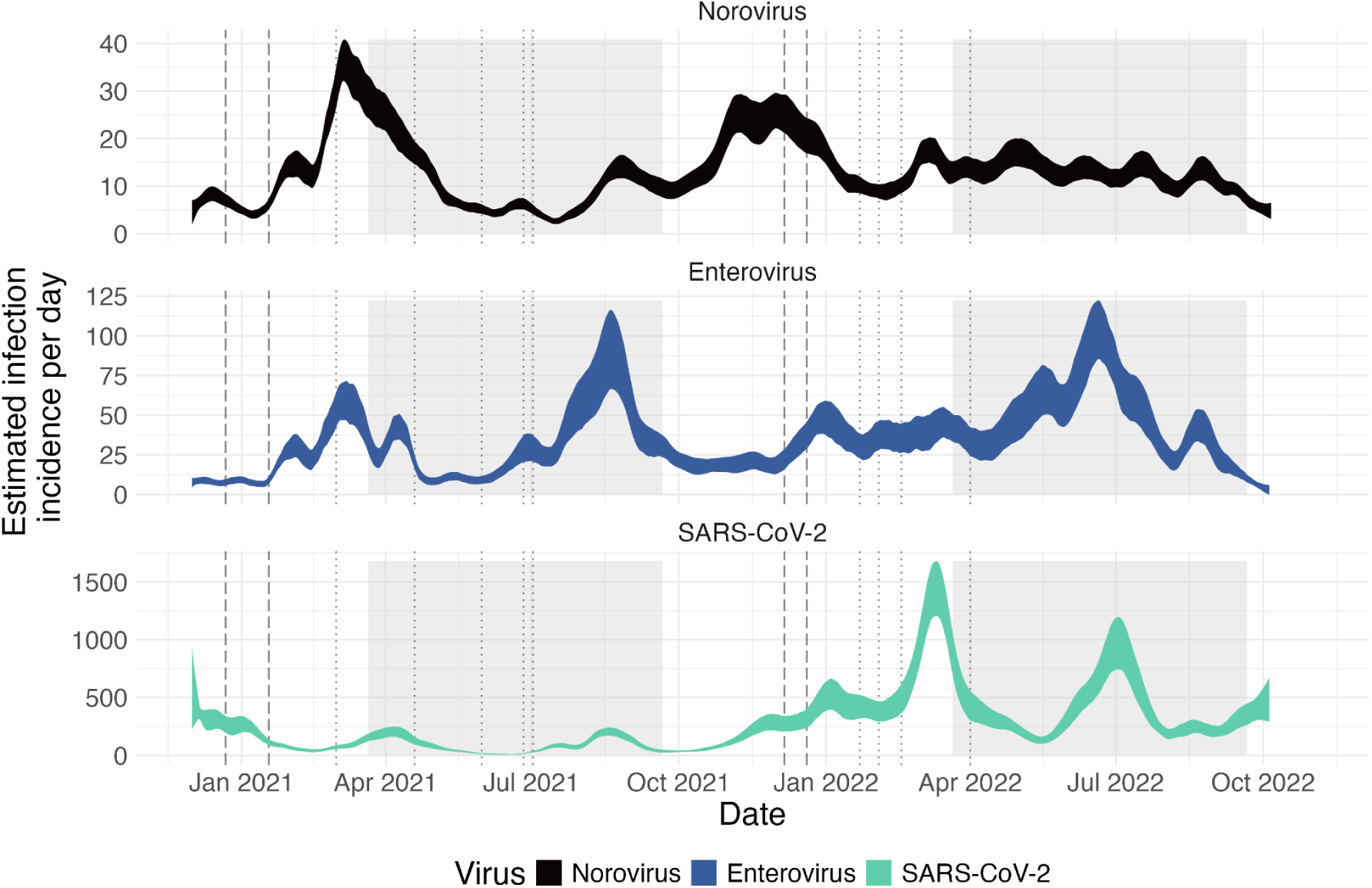
Estimated case incidence for NoV (top, black), EV (middle, blue) and SARS-CoV-2 (bottom, green). Grey shaded regions indicate the spring and summer months, white regions indicate fall and winter months. Dashed vertical lines indicate increases in COVID-19 containment measures, and dotted lines reductions in

**Figure S6:**
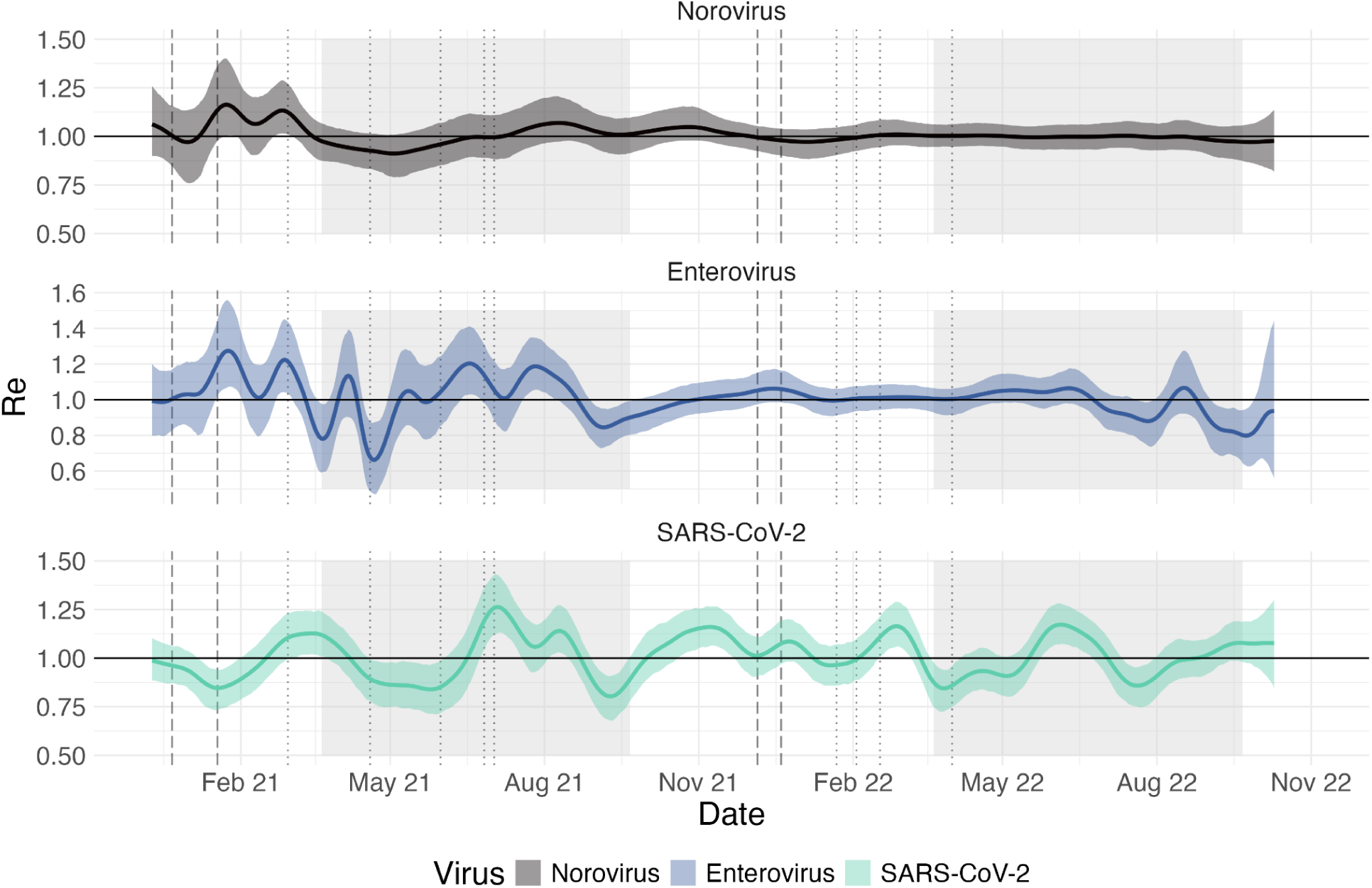
Estimated Re for NoV (top, black), EV (middle, blue), and SARS-CoV-2 (bottom, green) using the EpiSewer pipeline. Grey shaded regions indicate the spring and summer months, white regions indicate fall and winter months. Dashed vertical lines indicate increases in COVID-19 containment measures, and dotted lines reductions in stringency.

**Figure S7:**
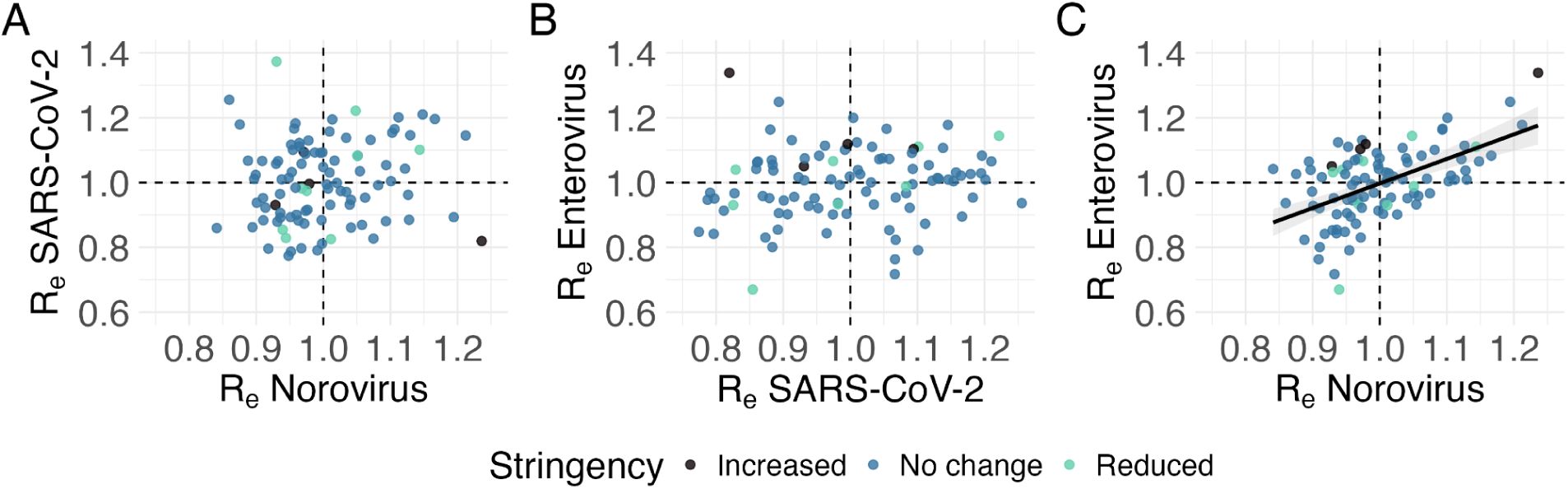
Correlation of the average Re per week between the three viruses. Points are colored by the change in KOF stringency index observed that week. The NoV and EV Re are significantly correlated (p-value: 6.3e-09).

## Supplementary Tables

**Table S1:** Changes in the KOF stringency index.

| **Date** | **Previous KOF value** | **Change in KOF value** | **Change in measures** |
| --- | --- | --- | --- |
| 2020-12-22 | 59.2 | +10.8 | Closure of restaurants, gyms, cultural institutions |
| 2021-01-18 | 70 | +5 | Closure of nonessential shops |
| 2021-03-01 | 75 | -10.8 | Lessened restrictions on shops, museums, zoos |
| 2021-04-19 | 64.2 | -8.33 | Lessened restrictions on public events and schools |
| 2021-05-31 | 55.8 | -10.8 | Lessened restrictions on gatherings and facial coverings |
| 2021-06-26 | 45 | -10 | Relaxed stay at home requirements |
| 2021-07-02 | 35 | -5 | Relaxation of public transport related restrictions |
| 2021-12-06 | 30 | +2.5 | Work from home recommendation |
| 2021-12-20 | 32.5 | +10 | Work from home mandate<br>Restrictions on gatherings + mandatory masking in public transport |
| 2022-01-22 | 42.5 | -2.5 | Vaccination or negative test required to enter Switzerland |
| 2022-02-03 | 40 | -7.5 | Lifting of work from home mandate |
| 2022-02-17 | 32.5 | -17.5 | Lifting of work from home recommendation and restrictions on the size of gatherings |
| 2022-04-01 | 15 | -15 | End of masking in public transport and isolation with positive test |

**Table S2:** Quarterly estimates of the effective reproductive number of NoV, EV, and SARS-CoV-2 respectively as estimated using EpiSewer.

|  | Q1<br>2021 | Q2<br>2021 | Q3<br>2021 | Q4<br>2021 | Q1<br>2022 | Q2<br>2022 | Q3<br>2022 |
| --- | --- | --- | --- | --- | --- | --- | --- |
| **Norovirus GII**<br>(mean ± sd) | 1.06 ±<br>0.06 | 0.95 ±<br>0.03 | 1.03 ±<br>0.02 | 1.02 ±<br>0.03 | 1.00 ±<br>0.01 | 1.00 ±<br>0.003 | 0.99 ±<br>0.01 |
| range [min, max] | [0.95,<br>1.16] | [0.91,<br>0.99] | [0.99,<br>1.07] | [0.97,<br>1.05] | [0.97,<br>1.01] | [1.00,<br>1.00] | [0.97,<br>1.00] |
| **Enterovirus**<br>(mean ± sd) | 1.07 ±<br>0.14 | 1.01 ±<br>0.16 | 1.01 ±<br>0.12 | 1.01 ±<br>0.04 | 1.01 ±<br>0.01 | 1.04 ±<br>0.02 | 0.91 ±<br>0.08 |
| range [min, max] | [0.78,<br>1.27] | [0.66,<br>1.20] | [0.85,<br>1.19] | [0.93,<br>1.06] | [1.00,<br>1.03] | [0.96,<br>1.06] | [0.80,<br>1.07] |
| **SARS-CoV-2**<br>(mean ± sd) | 0.99 ±<br>0.11 | 0.93 ±<br>0.11 | 1.03 ±<br>0.14 | 1.09 ±<br>0.05 | 1.01 ±<br>0.09 | 1.02 ±<br>0.11 | 0.98 ±<br>0.07 |
| range [min, max] | [0.85,<br>1.23] | [0.84,<br>1.27] | [0.80,<br>1.26] | [1.00,<br>1.16] | [0.84,<br>1.16] | [0.86,<br>1.17] | [0.86,<br>1.08] |

